## Supplementary Information for "Modelling the impact of interventions on imported, introduced and indigenous malaria infections in Zanzibar, Tanzania"

### S1 Methods

#### S1.1 Modelling of interventions

##### S1.1.1 RCD at a range of levels of follow up

The percentage of malaria cases diagnosed at a health facility that are followed up by a District Malaria Surveillance Officer is given by $\eta$. $\eta$ is varied between 0% and 100% to model the extreme values of removing RCD altogether, and perfect follow up of all cases diagnosed at a health facility.

##### S1.1.2 RCD at a range of levels of treatment seeking

The rate at which people seek treatment is $2.9 \times 10^{-4}$ per day in Pemba and $6.1 \times 10^{-4}$ per day in Unguja. This was calculated by considering the median number of malaria infections diagnosed at a health facility per month per district on Pemba and Unguja, and scaling by the number of districts and 30 days in a month [1]. This was increased by a factor of 2 or 3 as to simulate increase in treatment as an intervention.

##### S1.1.3 RCD with follow up of neighbours

Currently, neighbours are not included in RCD. We simulated the testing and treatment of 20 and 100 neighbours, as well as the index household. From the RADZEC study data, we estimated

*Corresponding author
†Current affiliations: Amsterdam Institute for Global Health and Development, Amsterdam, Netherlands and Amsterdam University Medical Centers, Amsterdam, Netherlands
‡Current affiliation: RTI International, Dar es Salaam, United Republic of Tanzania

that the targetting ratio amongst neighbouring households is around 0.7 (95% confidence interval (CI): 0.4–1.3) in Pemba and 1.3 (95% CI: 0.9–1.9) in Unguja.

Thus, the RCD term was modified to

$$\varphi_k = \rho I_k(\tau_k^{(h)}\nu_k^{(h)} + \tau_k^{(n)}\nu_k^{(n)})\eta\xi_k, \tag{1}$$

with $\nu_k^{(n)}$ either 20 or 100.

###### S1.1.4 Switching from RCD to RDA

When modelling RDA, we considered that all index household members and neighbours (when included) would receive treatment regardless of disease status. Thus, the diagnostic test sensitivity, $\rho$, was changed from 34% to 100%.

###### S1.1.5 Treatment of a proportion of cases brought on to Zanzibar by travelling humans (either residents or visitors)

Currently prophylaxis is not provided to travellers to mainland Tanzania. Similarly, there is no screen-and-treat programme for entrants to Zanzibar. We include treatment of imported cases as a potential intervention in our model, in order to evaluate what proportion of cases must be treated to achieve different reductions in prevalence on Pemba and Unguja [2]. We modify Eq. (4) in the main text to have a $\theta^{\text{outbound}}$, which includes treatment for visitors from mainland Tanzania on their outbound journey to Zanzibar, and $\theta^{\text{return}}$ for Zanzibari residents that receive treatment on their return journey to Zanzibar. Thus the base form of the equation becomes

$$\frac{\mathrm{d}I_k}{\mathrm{d}t} = \sum_{i=1}^{3}\left(\beta_i\left(\frac{\sum_{j=1}^{3} N_j\theta_{ij}^{\text{outbound}} I_j}{\sum_{j=1}^{3} N_j\theta_{ij}}\right)\theta_{ik}^{\text{return}}\right)(1 - I_k) - (\mu + \varphi_k)I_k, \tag{2}$$

where

$$\theta^{\text{outbound}} = \begin{pmatrix} 0.991 & 0.004 & (1-O)*5.7\times10^{-5} \\ 0.003 & 0.970 & (1-O)*5.3\times10^{-4} \\ 0.006 & 0.026 & 0.999 \end{pmatrix}, \tag{3}$$

and

$$\theta^{\text{return}} = \begin{pmatrix} 0.991 & 0.004 & 5.7\times10^{-5} \\ 0.003 & 0.970 & 5.3\times10^{-4} \\ (1-R)*0.006 & (1-R)*0.026 & 0.999 \end{pmatrix}. \tag{4}$$

$O$ represents the proportion of travellers from mainland Tanzania receiving treatment such that they are no longer infected upon entering Zanzibar, and $R$ represents the proportion of Zanzibari residents receiving treatment such that they are no longer infected upon returning to Zanzibar. We always simulate equal proportions of outbound and return cases being treated (i.e. $O = R$)

###### S1.1.6 Reductions in the malaria transmission rate

The rate at which malaria is transmitted from one human to another can be reduced through vector control interventions such as the use of long-lasting insecticidal nets, indoor residual spraying and larval source management. As we do not explicitly model mosquitoes or the vectorial capacity, we reduce the transmission parameter, $\beta$, to simulate increases in vector control.

Thus, for this intervention, $\beta$ is replaced by $\beta(1-r)$ where $r$ is the reduction in vectorial capacity. As vectorial capacity is proportional to the number of susceptible humans infected by an infected human per day, $\beta$ is proportional to the vectorial capacity, and any reduction in $\beta$ could arise from a proportional reduction in the vectorial capacity [3]. Values ranging from 0.25 to 0.9 were tested for $r$ on Pemba and Unguja, and values ranging from 0.1 to 0.3 for $r$ on mainland Tanzania.

### S2 Results

#### S2.1 Comparison of interventions

The impact of each intervention alone was tested by changing one factor at a time and plotting the final equilibrium reached 40 years after the introduction of the intervention. All other factors were held at their baseline value, given in Table 3 in the main text. The results from this analysis are shown in Fig S1. Most intervention parameters had an approximately linear relationship with malaria incidence, but the relationship between the percentage of travellers treated and the incidence of infections was mildly concave, and the relationship between a reduction in the malaria transmission rate in Zanzibar and the incidence of infections was steeply curved. This suggests that even small increases in vector control may have a disproportionately large impact with regards to reducing malaria incidence on Zanzibar.

#### S2.2 Impact of parameter uncertainty

Parameter uncertainty was considered in the same way as described in Das *et al* (2022) [2]. Simulations were run with a range of parameter values based on the uncertainty in the data. The parameters varied and the distributions from which they were sampled were as follows:

- The equilibrium malaria prevalence on Pemba, $I_1^* \sim \text{Beta}(32, 2242)$ ;
- The equilibrium malaria prevalence on Unguja, $I_2^* \sim \text{Beta}(92, 3196)$;
- The targeting ratio in index households in Pemba, $\tau_1^{(h)} \sim \frac{\text{Beta}(20,427)}{I_1^*}$;
- The targeting ratio in index households in Unguja, $\tau_2^{(h)} \sim \frac{\text{Beta}(64,470)}{I_2^*}$;
- The targeting ratio in neighbouring households in Pemba, $\tau_1^{(n)} \sim \frac{\text{Beta}(13,1147)}{I_1^*}$;
- The targeting ratio in neighbouring households in Unguja, $\tau_2^{(n)} \sim \frac{\text{Beta}(26,1619)}{I_2^*}$;
- The number of people tested by the RCD programme in the index household in Pemba, $\nu_1^{(h)} \sim \text{Normal}(7.02, 0.24)$;
- The number of people tested by the RCD programme in the index household in Unguja, $\nu_2^{(h)} \sim \text{Normal}(6.36, 0.25)$;
- The number of people tested by the RCD programme in neighbouring households in Pemba, $\nu_1^{(n)} \sim \text{Normal}(20.36, 0.50)$;

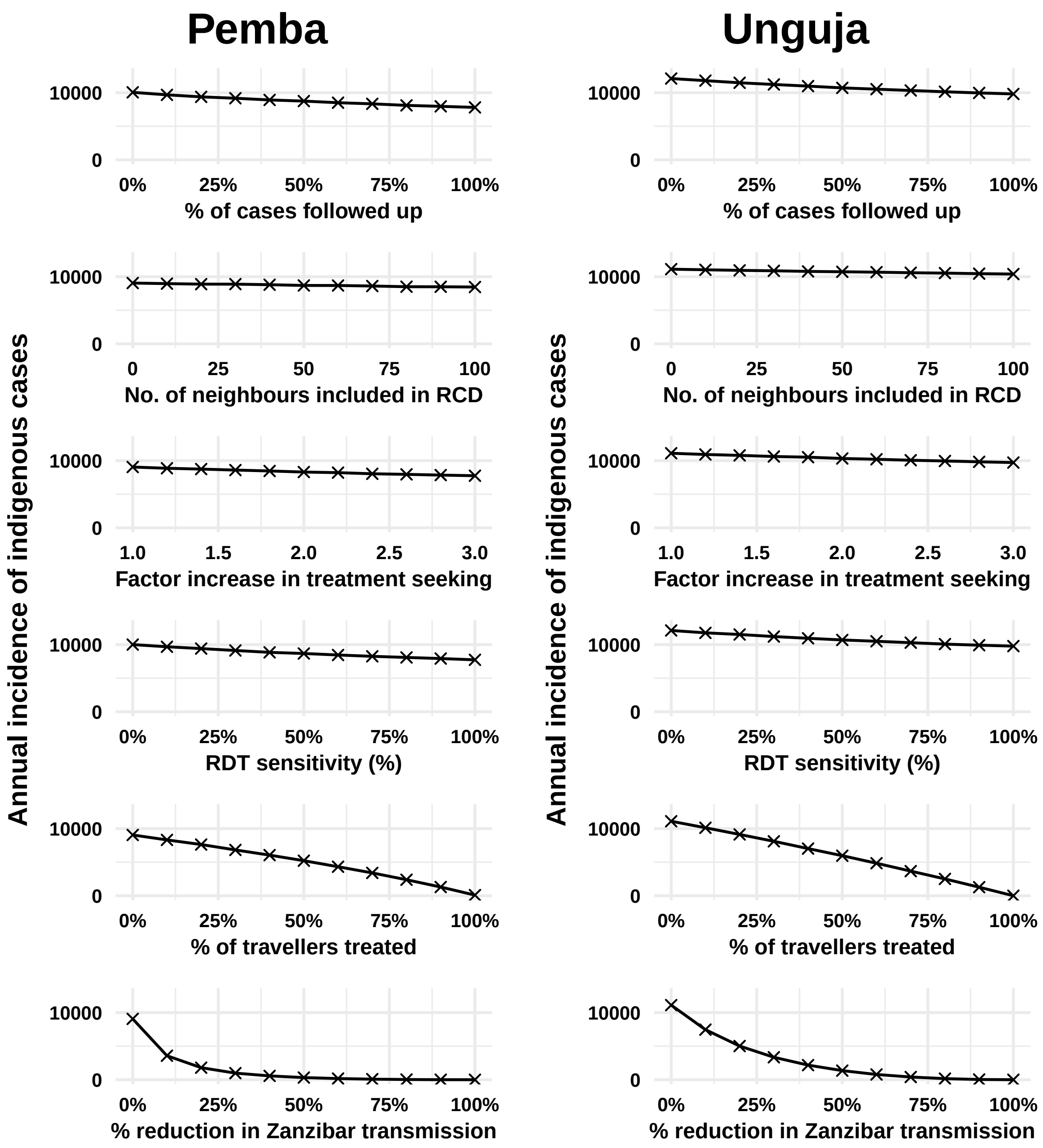


**Figure S1:** Median yearly incidence of indigenous cases out of 500 simulations in the 40$^{\text{th}}$ year after the start of each intervention. At each point, only the parameter on the x-axis has been changed, with all other parameters remaining at the baseline value. RCD: reactive case detection; RDT: rapid diagnostic test.

- The number of people tested by the RCD programme in neighbouring households in Unguja, $\nu_2^{(n)} \sim \text{Normal}(18.76, 0.58)$.

Subscripts of *1* and *2* indicate Pemba and Unguja, respectively. Parameter values with the 95% interval values can be found in Table S1.

100 random values were selected from these parameter distributions, and each set of values was simulated with five different seeds, forming a total of 500 simulations for each intervention scenario. The final equilibrium value reached for a range of interventions, along with the uncertainty stemming from both the parameter and stochastic variation, is shown in Fig S2. The

| Variable or parameter | Mean values [95% CI] | | | Source |
|---|---|---|---|---|
| | Pemba | Unguja | Mainland | |
| $I_k^*$ | 1.36% [0.96-1.93] | 1.18% [0.86-1.61] | 7.79% | [4, 5] |
| $N_k$ | 406,848 | 896,721 | 43,625,354 | [6] |
| $\theta_{ij}$ | $\begin{array}{r} \\ \text{Pemba} \\ \text{Unguja} \\ \text{Mainland} \end{array} \begin{array}{c} \begin{array}{ccc} \text{Pemba} & \text{Unguja} & \text{Mainland} \end{array} \\ \begin{pmatrix} 0.991 & 0.004 & 5.7\times10^{-5} \\ 0.003 & 0.970 & 5.3\times10^{-4} \\ 0.006 & 0.026 & 0.999 \end{pmatrix} \end{array}$ | | | [4] |
| $\mu$ | 0.005 $\text{day}^{-1}$ | 0.005 $\text{day}^{-1}$ | 0.005 $\text{day}^{-1}$ | [7, 8] |
| $\tau_k^{(h)}$ | 3.2 [2.0-4.8] | 10.0 [8.0-12.6] | N/A | [4] |
| $\tau_k^{(n)}$ | 0.7 [0.4-1.3] | 1.3 [0.9-1.9] | N/A | [4] |
| $\nu_k^{(h)}$ | 7.0 [6.5-7.5] | 6.3 [5.9-6.9] | N/A | [4] |
| $\nu_k^{(n)}$ | 20.4 [19.4-21.4] | 18.8 [17.6-19.9] | N/A | [4] |
| $\rho^*$ | 34% | 34% | N/A | [4] |
| $\eta^*$ | 35.3% | 35.3% | N/A | [1] |
| $\xi^*$ | $2.9 \times 10^{-4}$ $\text{day}^{-1}$ | $6.1 \times 10^{-4}$ $\text{day}^{-1}$ | N/A | [4, 1] |

**Table S1:** Variable and parameter values at baseline and sources. Where a range of parameter values were tested in the uncertainty analysis, the 95% confidence interval for the range of values tested is given. The superscripts $(h)$ and $(n)$ indicate the index household and neighbouring households, respectively.

impact of parameter uncertainty on the probability of reaching elimination was also examined and found to be minor (see Fig S3). Even when parameter uncertainty is included, elimination is only observed when there is 100% importation treatment in the absence of transmission reduction interventions.

#### S2.3 Sensitivity analysis

A sensitivity analysis was conducted using the Sobol method to characterise the impact of parameter variation on model outputs. 32,768 parameter values were sampled from uniform distributions for each parameter using Saltelli sampling [9, 10]. The bounds of the uniform distributions corresponded to 95% confidence intervals found in the literature or, when such bounds were not available, the point estimate of the parameter ±50%. Upper and lower bounds and data sources can be found in Table S2. The upper bound of the proportion of time spent by mainland Tanzania residents on Pemba and Unguja was calculated by scaling the upper bound from Le Menach *et al* (2011) by the proportion of people residing on mainland Tanzania as compared to Pemba or Unguja. The model was then run using these parameter sets and a different seed for each parameter set, and the outputs were used to calculate Sobol indices. First order and total Sobol indices were calculated using the SALib package (version 1.4.5) in Python [10].

This analysis suggests that the main outputs of malaria prevalence and the annual incidence of indigenous infections are most sensitive to the estimates of the transmission parameter (Fig S4 and S5). As these are back-calculated from the baseline malaria prevalence, the need for accurate estimates of the prevalence in the general population is important for a correct estimate of

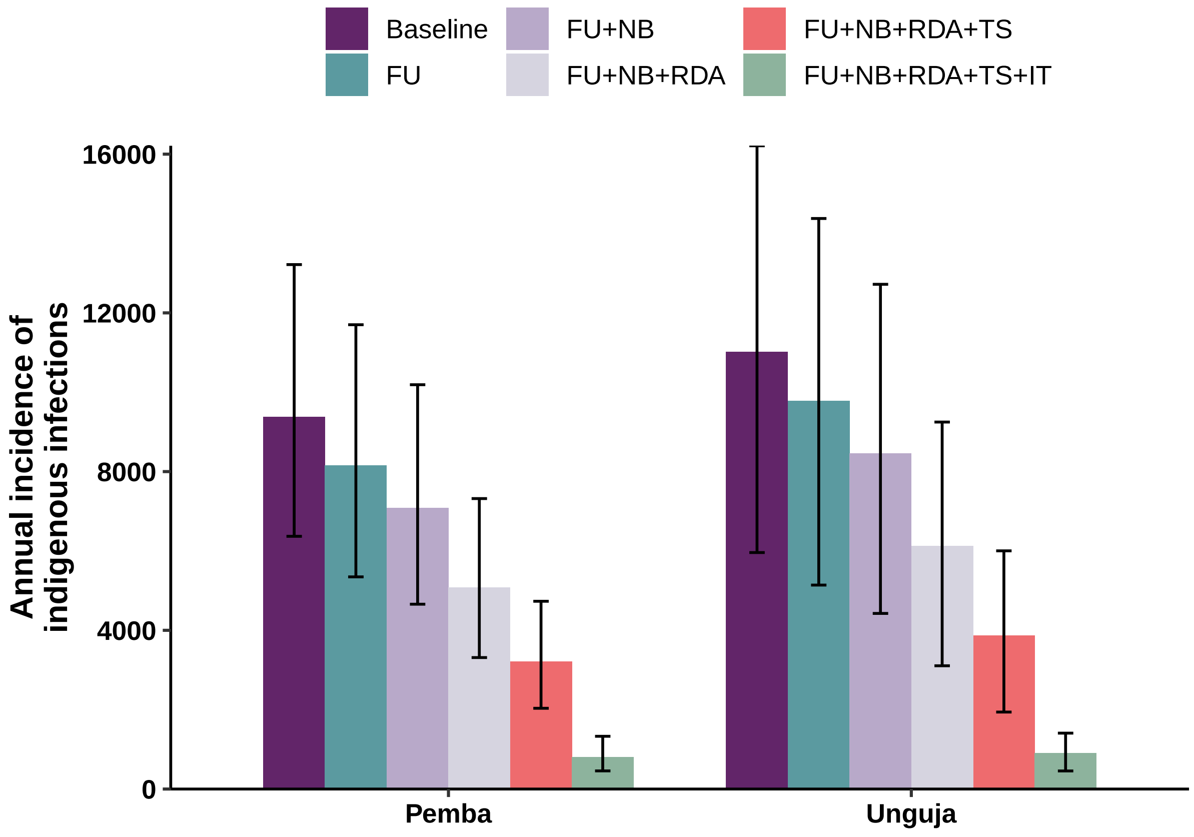


**Figure S2:** Bar chart showing the median yearly incidence of indigenous infections 40 years after the start of interventions (i.e. once equilibrium is reached). The error bars indicate the 95% confidence for both the parameter and stochastic uncertainty. FU: 100% follow up; NB: 100 neighbours included in testing and treatment; RDA: reactive drug administration; TS: three times the baseline treatment seeking rate; IT: treatment of 90% of travellers arriving on Zanzibar.

the effective human-to-human malaria transmission rate. An accurate estimate of the baseline prevalence is also key to estimating the probability of elimination being reached under different intervention conditions.

#### S2.4 The impact of a fixed versus a varying targetting ratio

The targetting ratio is calculated from the RADZEC study data and is assumed to be fixed in the main text, regardless of the population malaria prevalence. This implies that cases do not become more clustered as the disease prevalence falls. In comparison, Chitnis *et al* (2019) consider a targetting ratio that varies depending on prevalence and the number of people tested, with the ratio of malaria infections amongst those tested as compared to the general population decreasing as prevalence and the number of people tested increases [13]. The following function was found to best estimate the targetting ratio, $\tau$, for geo-located prevalence data collected in Zambia:

$$\tau(\nu, I) = \exp\left((-\alpha_1 \ln(I) + \frac{\alpha_2}{\nu} - \frac{\alpha_3}{\nu}\ln(I))\frac{N-\nu}{N}\right) \mid \tau(\nu, I) \geq 1, \tag{5}$$

where $\tau$ is the targetting ratio, $\nu$ is the number of people tested, not including the index case, $N$ is the total population, and $\alpha_1$, $\alpha_2$ and $\alpha_3$ are fitted parameters with values $\alpha_1 = 0.23$

| **Parameter** | **Point estimate** | **Lower bound** | **Upper bound** | **Reference** |
|---|---|---|---|---|
| Transmission rate on Pemba ($\beta_{\text{Pemba}}$) | 0.0048 | 0.0024 | 0.0072 | ±50% |
| Transmission rate on Unguja ($\beta_{\text{Unguja}}$) | 0.0037 | 0.0019 | 0.0056 | ±50% |
| Transmission rate on mainland Tanzania ($\beta_{\text{Mainland}}$) | 0.0054 | 0.0027 | 0.0081 | ±50% |
| Mean duration of infection ($1/\mu$) | 200 | 184 | 237 | [11] |
| Targeting ratio on Pemba ($\tau^{(h)}_{\text{Pemba}}$) | 3.1 | 2.0 | 4.8 | RADZEC data ([4]) |
| Targeting ratio on Unguja ($\tau^{(h)}_{\text{Unguja}}$) | 10.1 | 8.0 | 12.7 | RADZEC data ([4]) |
| Treatment seeking rate on Pemba ($\xi_{\text{Pemba}}$) | $2.9 \times 10^{-4}$ | $1.5 \times 10^{-4}$ | $4.4 \times 10^{-4}$ | ±50% |
| Treatment seeking rate on Unguja ($\xi_{\text{Unguja}}$) | $6.1 \times 10^{-4}$ | $3.1 \times 10^{-4}$ | $9.2 \times 10^{-4}$ | ±50% |
| RDT sensitivity ($\rho$) | 34% | 0% | 100% | Full range |
| Follow up of index cases ($\eta$) | 35% | 0% | 100% | Full range |
| Movement from Pemba to Unguja ($\theta_{UP}$) | 0.0032 | 0.0016 | 0.0048 | ±50% |
| Movement from Pemba to mainland Tanzania ($\theta_{MP}$) | 0.0061 | 0 | 0.12 | [12] |
| Movement from Unguja to Pemba ($\theta_{PU}$) | 0.0039 | 0.0019 | 0.0058 | ±50% |
| Movement from Unguja to mainland Tanzania ($\theta_{MU}$) | 0.026 | 0 | 0.12 | [12] |
| Movement from mainland Tanzania to Pemba ($\theta_{PM}$) | $5.7 \times 10^{-5}$ | 0 | 0.0011 | [12] |
| Movement from mainland Tanzania to Unguja ($\theta_{UM}$) | $5.3 \times 10^{-4}$ | 0 | 0.0025 | [12] |

**Table S2:** Parameter bounds for sensitivity analysis.

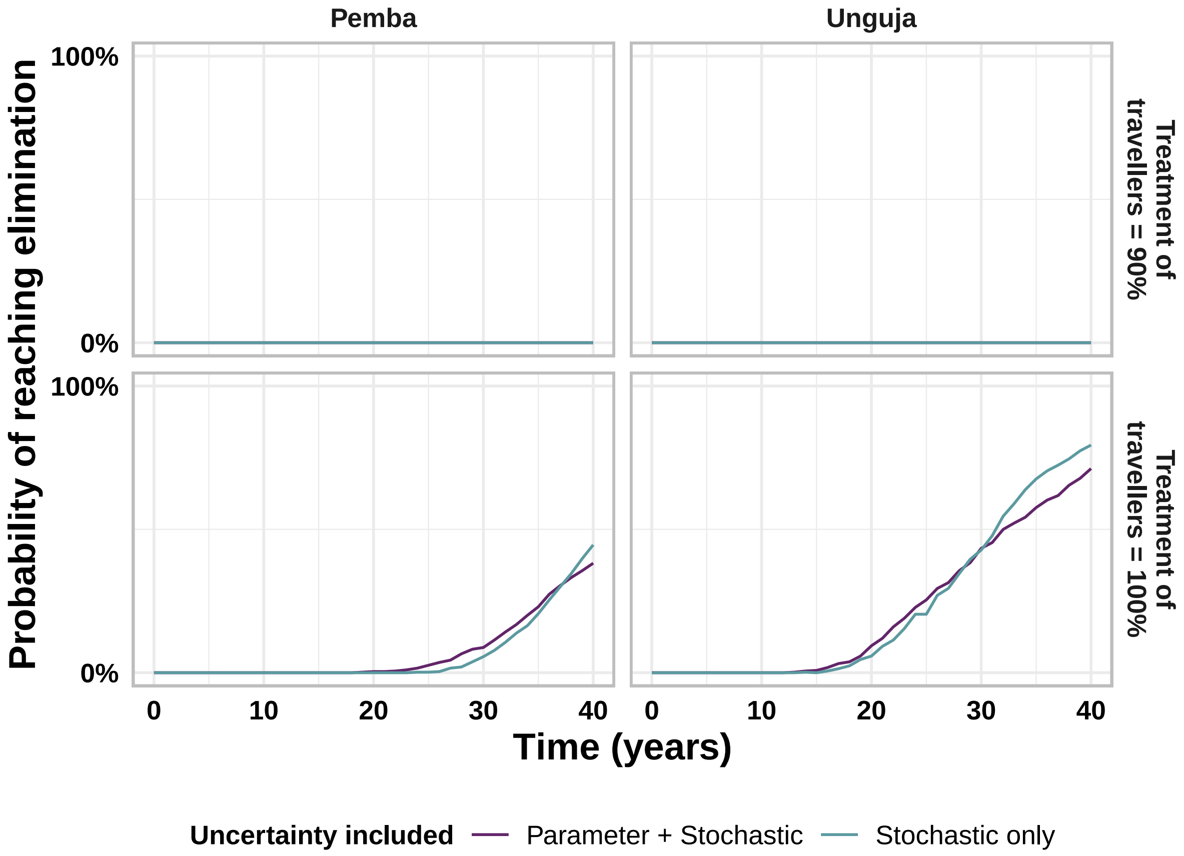


**Figure S3:** Proportion of stochastic simulations reaching elimination (three years with zero indigenous cases), starting from the introduction of all RCD-related interventions and treatment of imported cases, comparing stochastic uncertainty to the combination of parameter and stochastic uncertainty.

(95% credible interval (CI): 0.16, 0.29); $\alpha_2 = -1.40$ (CI: -2.77, -0.02) and $\alpha_3 = 2.87$ (CI: 1.13, 4.59) [13].

In order to compare running the model with a fixed targetting ratio and a varying targetting ratio, we take the function from Chitnis *et al* (2019) that is fitted to data from Zambia, and apply a scaling factor to adjust the targetting ratio so that the targetting ratio matches between Eq. (5) and the targetting ratio for the index household in the RADZEC data ($\tau^{(h)}$ in the main text). Thus, the equation we used to generate a varying $\tau$ was given by

$$\tau(\nu, I) = A \exp\left((-\alpha_1 \ln(I) + \frac{\alpha_2}{\nu} - \frac{\alpha_3}{\nu}\ln(I))\frac{N-\nu}{N}\right) \mid \tau(\nu, I) \geq 1, \tag{6}$$

where A was calculated to be 0.19 for Pemba and 0.42 for Unguja, in order to match the targetting ratios calculated by Eq. (6) and the targetting ratio seen in the RADZEC study data. Thus, as the malaria prevalence decreases due to the introduction of new interventions, the targetting ratio increases and the effectiveness of RCD increases.

Running the model with a varying $\tau$ and a fixed $\tau$, we see that the difference in the targetting ratio is not substantial even when considering the maximum RCD interventions tested i.e. RDA with triple the usual treatment seeking rate and 100 neighbours included in treatment (see Fig S6). These interventions maximise the effect of the targetting ratio and so are the ones where we'd expect to see the largest difference between the blue and purple lines in Fig S6. When RCD finds and treats a lot of cases, a targetting ratio that improves as the prevalence falls can

| | Number of imported cases | Number of infected visitors |
|---|---|---|
| **Pemba** | 216 | 234 |
| **Unguja** | 1888 | 1829 |

**Table S3:** Median number of imported cases and infected visitors present on each patch at equilibrium.

provide an optimistic outlook of potentially eliminating malaria earlier than when considering a fixed targetting ratio, which makes the more conservative assumption of no increase in case clustering as prevalence decreases. Nonetheless, given the difference is small, we have used a fixed targetting ratio for all simulations shown in the main text.

##### S2.5 The impact of varying the definition of malaria re-establishment

Currently, we consider a simulation to have reached elimination when three years have passed with zero incidence of indigenous cases. However, if an indigenous case appears after this three year period, we count this as malaria re-establishment and thus losing ‘eliminated’ status. In contrast, the World Health Organization defines the minimum indication of re-establishment of transmission as *‘the occurrence of three of more indigenous malaria cases of the same species per year in the same focus, for three consecutive years’* [14]. Since no country that has been certified as malaria-free has lost certification, we additionally modelled the impact of assuming that once a region eliminates malaria, it stays malaria-free. Out of the 500 simulations, when a simulation reaches three years with zero incidence of indigenous cases, we assume it remains at zero indigenous cases indefinitely into the future. The probability of reaching elimination is shown in Fig S7, with the assumption of remaining malaria-free after elimination labelled as ‘cumulative’ and the more strict definition of malaria re-establishment (losing ‘eliminated’ status after the appearance of one indigenous case) labelled as ‘transient’. We observe that in the majority of cases, the definition of re-establishment does not impact the proportion of runs reaching elimination. Only in the case where the number of indigenous cases is typically zero, but not always (90% importation treatment with 90% reduction in the transmission rate on Pemba) does the definition of re-establishment make a substantial difference to the number of runs reaching elimination.

### S3 Additional figures

#### S3.1 Time-series plots for individual interventions

#### S3.2 Figures with all previously introduced interventions also in place at maximum values

#### S3.3 Probability of elimination over a longer period of time with 100% treatment of travellers

### S4 Additional tables

[6] Tanzania National Bureau of Statistics. Population and housing census: population distribution by administrative areas. *Ministry of Finance, Dar es Salaam*, 2012.

[14] World Health Organization. *A framework for malaria elimination.* World Health Organization, 2017.

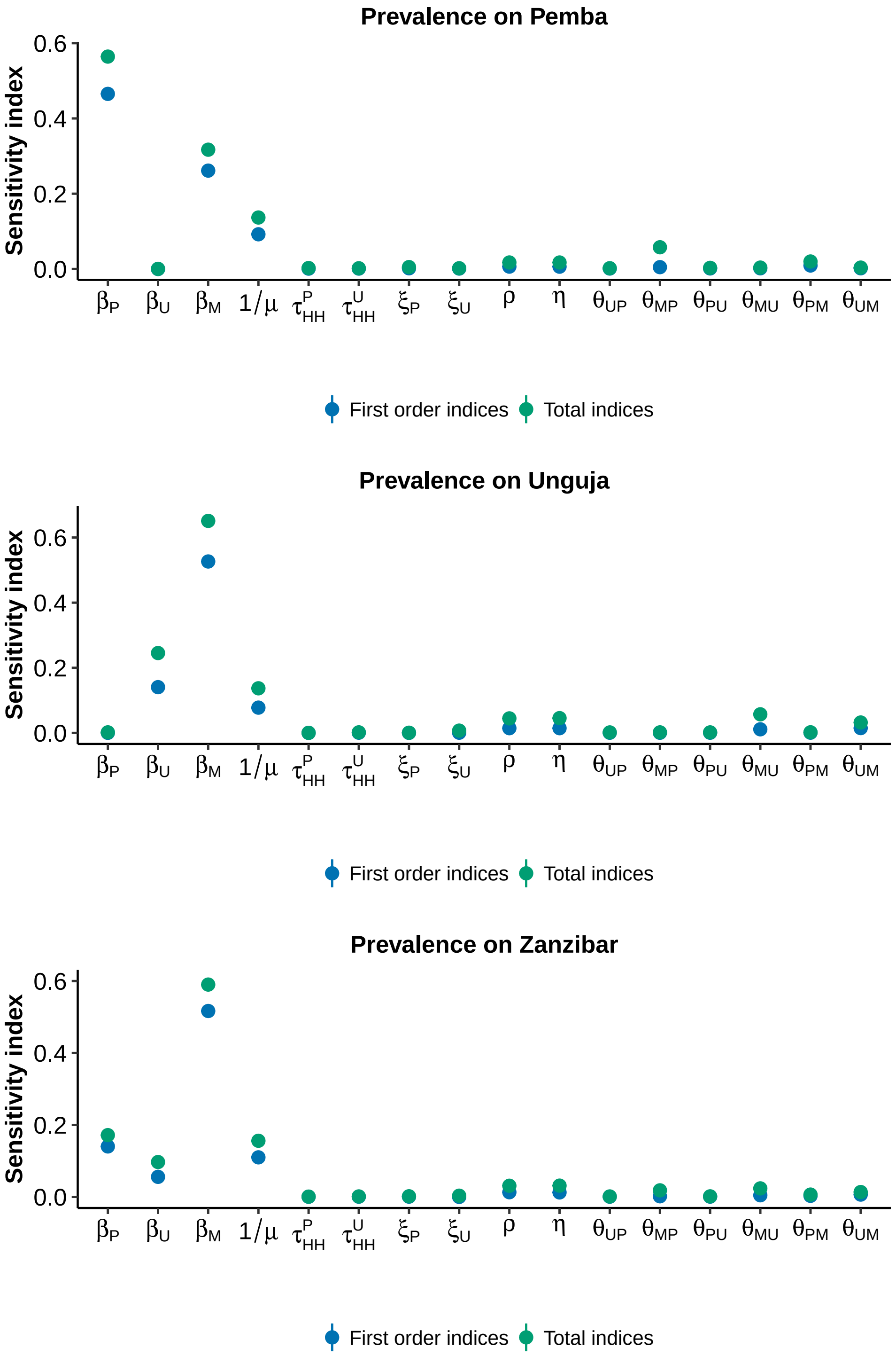


**Figure S4:** First order and total Sobol indices for each parameter tested when the overall malaria prevalence is considered as the output. Note, the 95% confidence intervals are smaller than the point sizes and so are not visible. The model output of malaria prevalence on Zanzibar as a whole was calculated by multiplying the expected prevalence on each island by the population size of each island, summing to get the total number of infected people on both islands, and then dividing by the summed population across both islands.

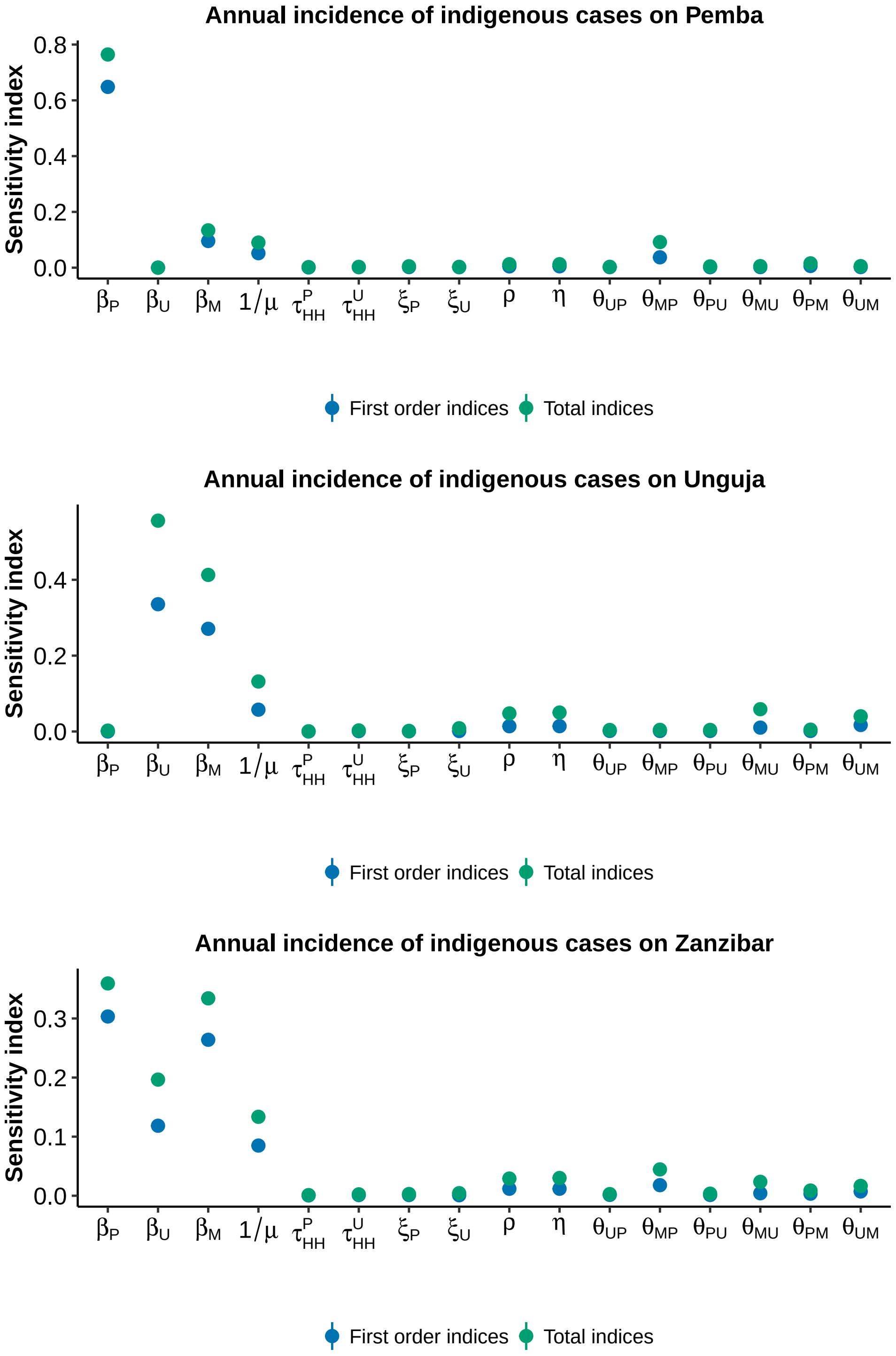


**Figure S5:** First order and total Sobol indices for each parameter tested when the annual incidence of indigenous cases is considered as the output. Note, the 95% confidence intervals are smaller than the point sizes and so are not visible. The total incidence of indigenous cases for Zanzibar as a whole was calculated by summing the incidence of indigenous cases on each island.

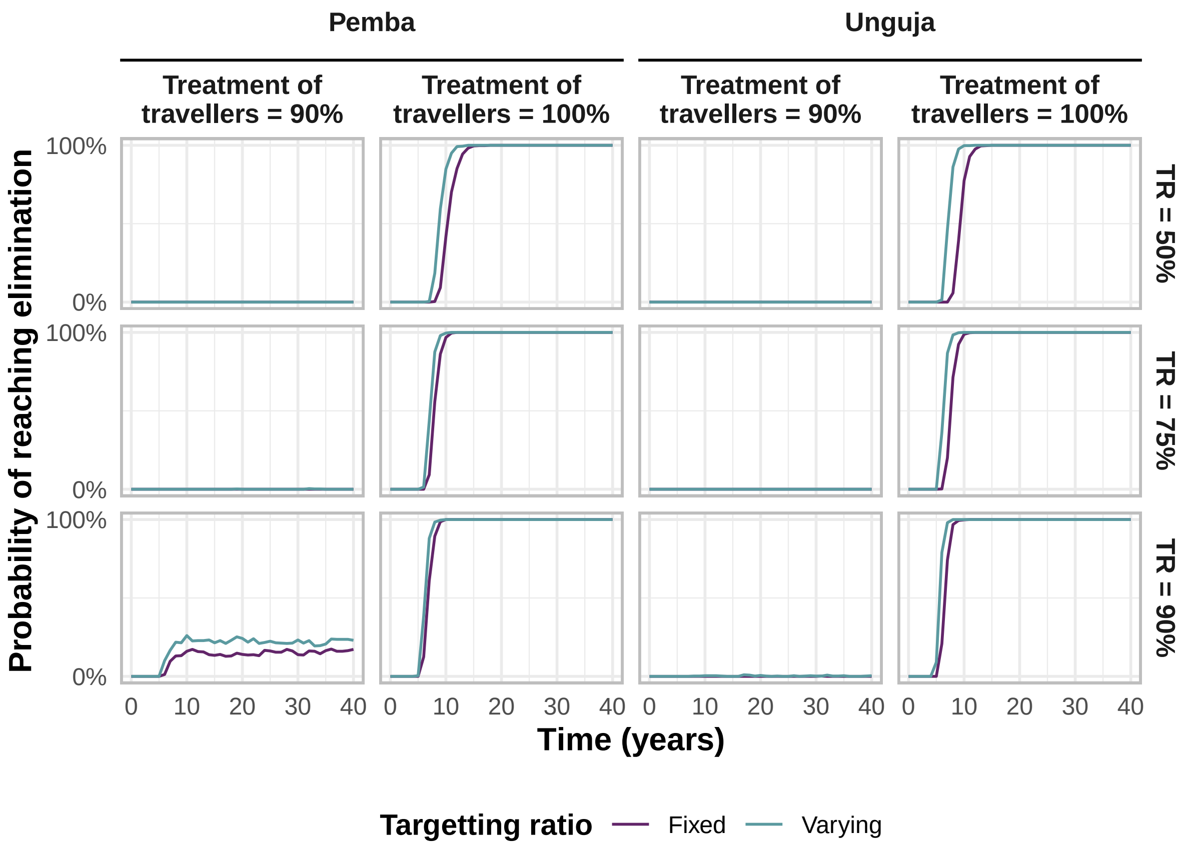


**Figure S6:** Proportion of stochastic simulations reaching elimination (three years with zero indigenous cases), starting from the introduction of all RCD-related interventions, comparing a more conservative definition of the targetting ratio, where the ratio is constant regardless of prevalence, and a definition where the targetting ratio increases as the malaria prevalence decreases.

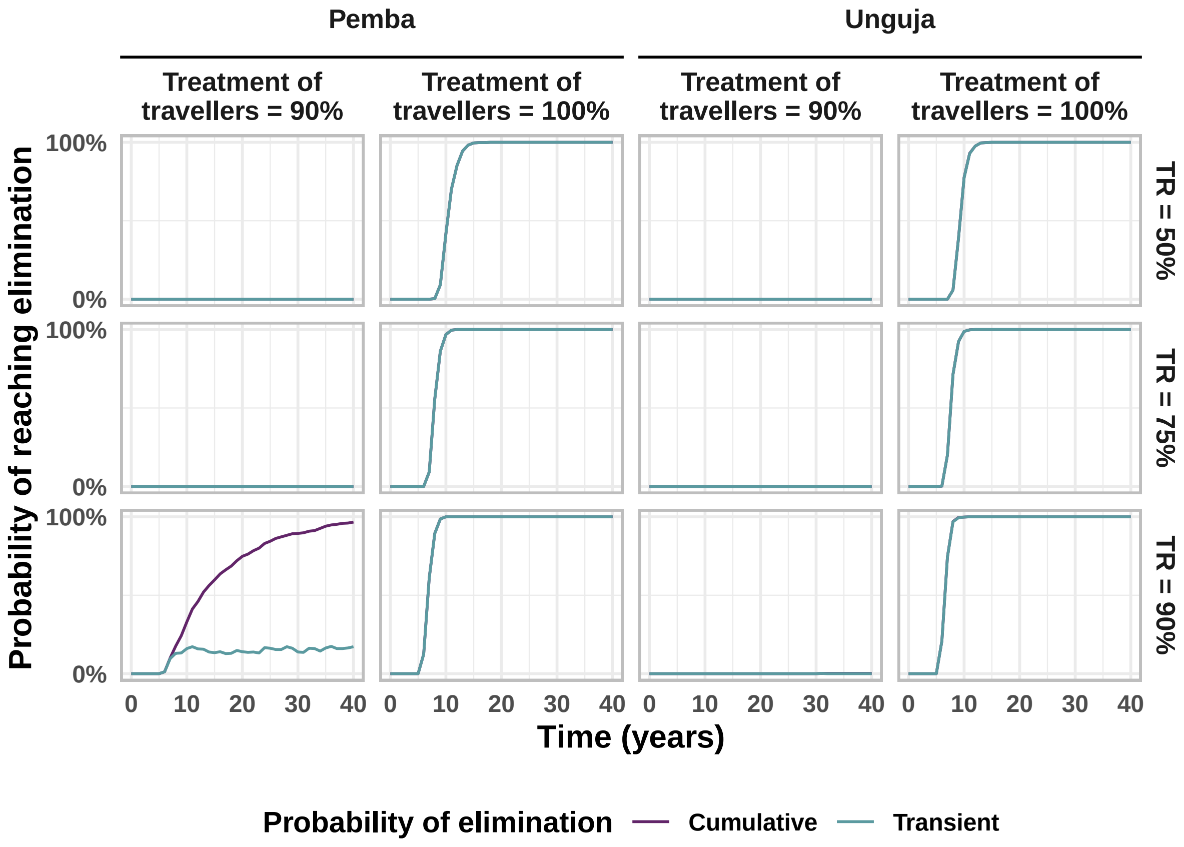


**Figure S7:** Proportion of stochastic simulations reaching elimination (three years with zero indigenous cases), starting from the introduction of all RCD-related interventions and treatment of imported cases, comparing a transient probability of elimination (where 'eliminated' status is lost after the appearance of one indigenous case), to a cumulative probability of elimination (once a simulation reaches elimination, it stays malaria-free with zero indigenous cases). 'TR' stands for transmission reduction.

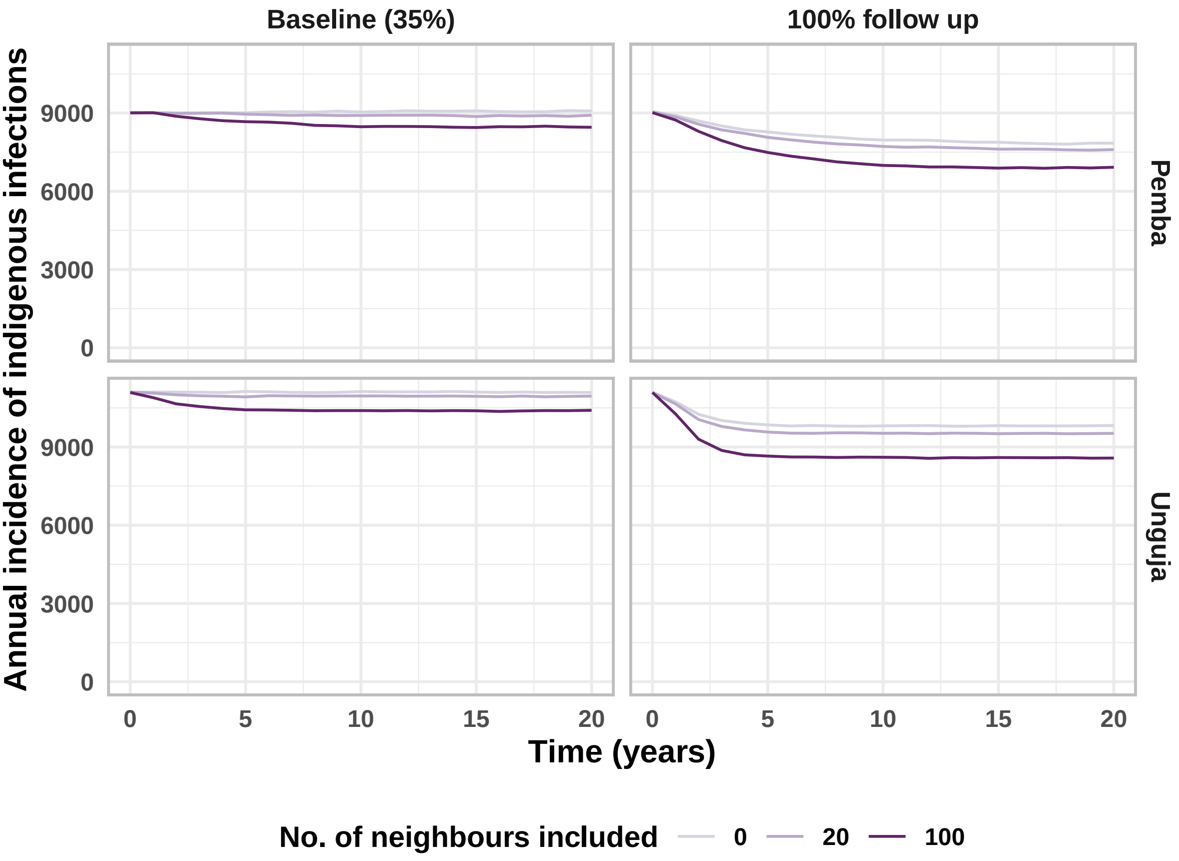


**Figure S8:** Median annual incidence of infections comparing the current RCD system to a system where 100% of malaria cases diagnosed at a health facility are followed up at the index household level and a range of number of neighbours are included in RCD.

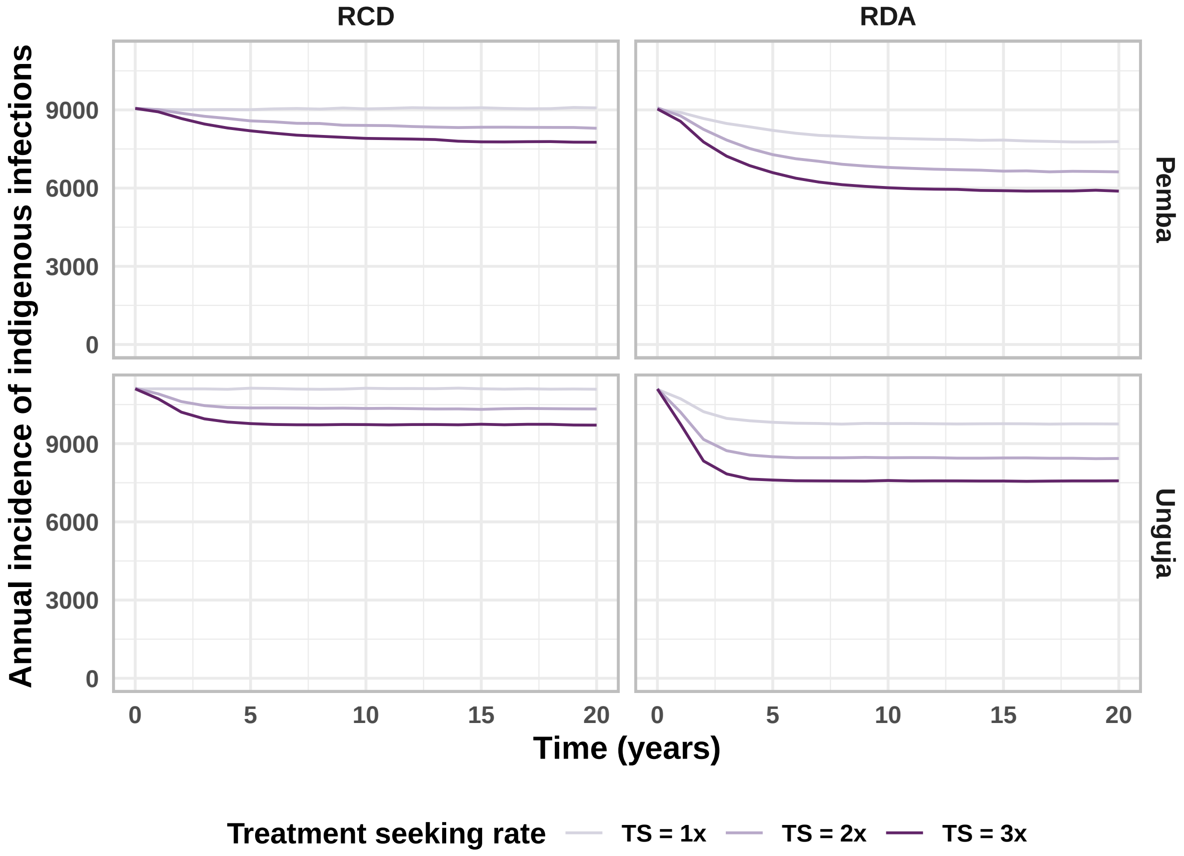


**Figure S9:** Median annual incidence of infections comparing the current RCD system to reactive drug administration (in reaction to detecting a case at a health facility, upon follow up, testing is skipped and antimalarials are given to all index household members) and increases in the rate at which people seek treatment (treatment seeking is abbreviated as 'TS').

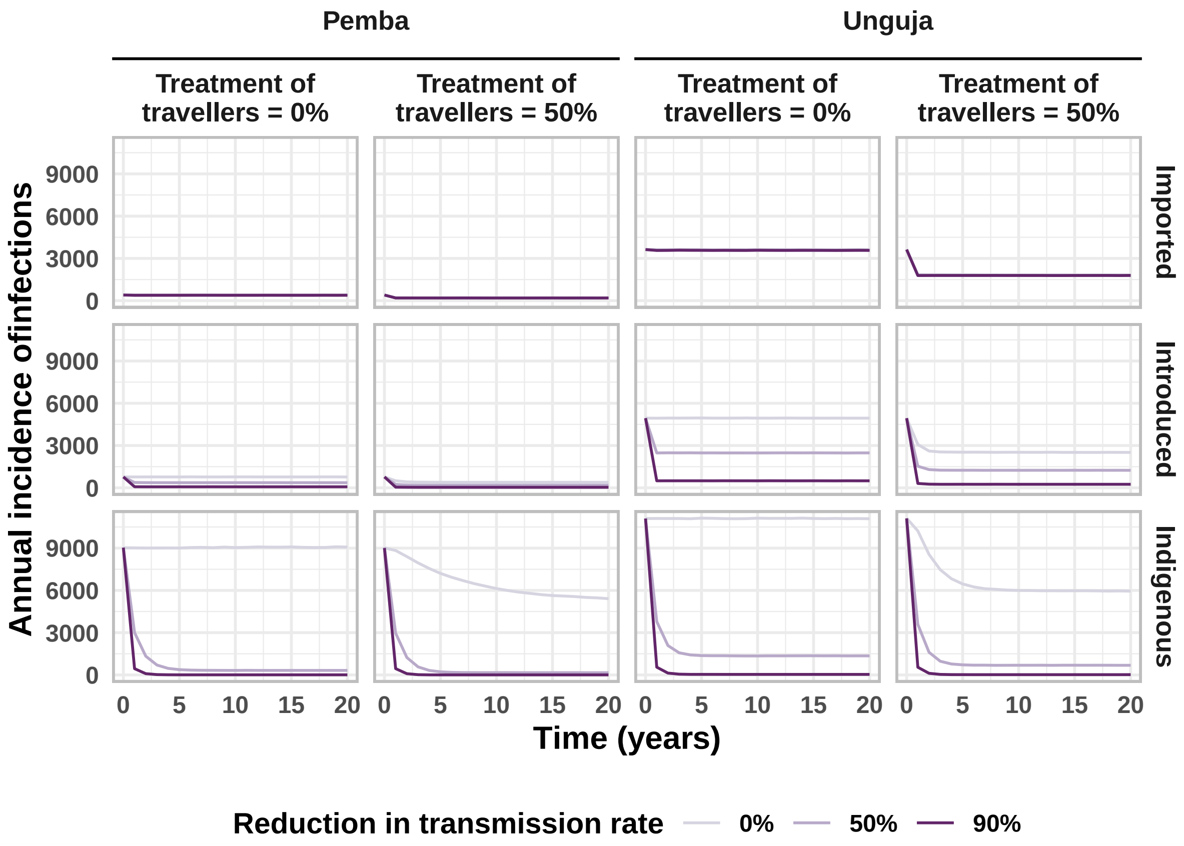


**Figure S10:** Median annual incidence of indigenous infections comparing the baseline interventions (RCD for 35% of cases arriving at a health facility at the index household level only) to also treating a range of proportions of infected travellers, as well as vector control to reduce the malaria transmission rate on Zanzibar.

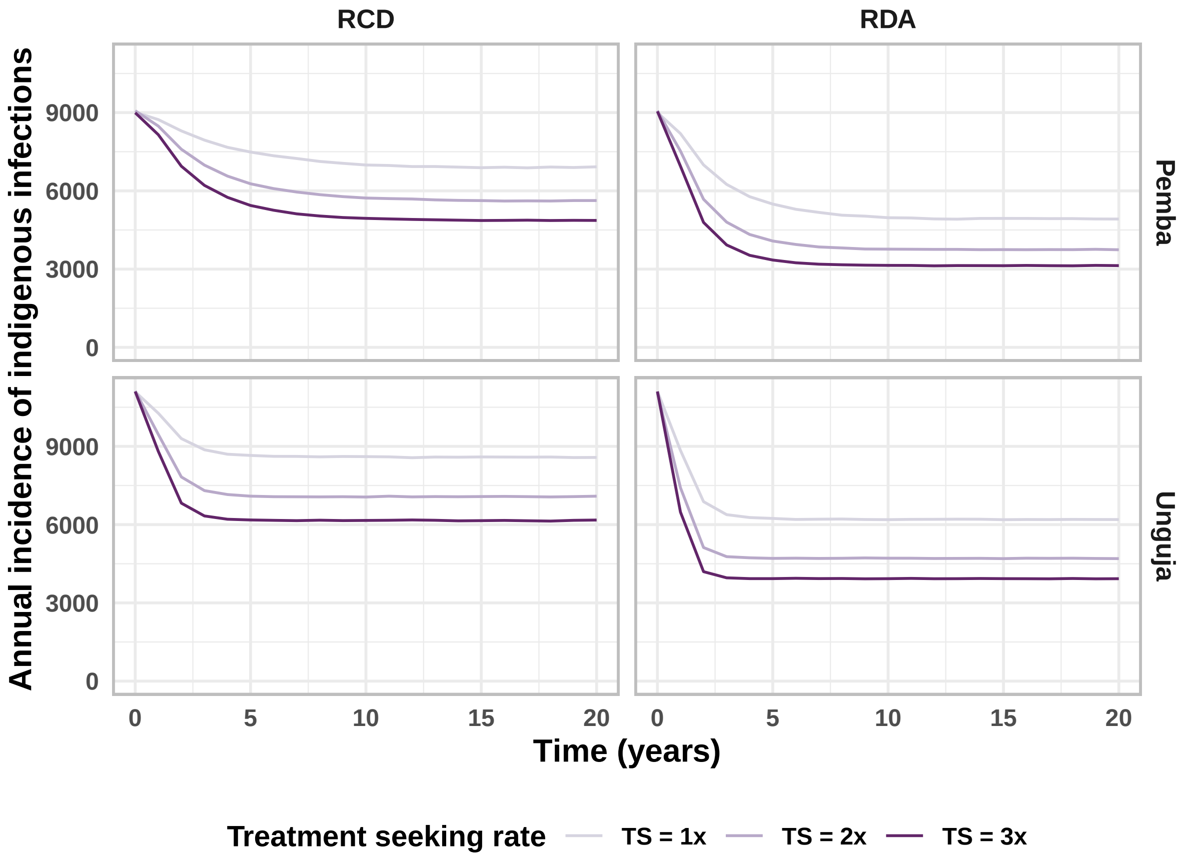


**Figure S11:** Median incidence of infections from 500 stochastic simulations comparing the current RCD system with 100% follow up of cases and 100 neighbours being included in testing and treating to reactive drug administration (in reaction to detecting a case at a health facility, upon follow up, testing is skipped and antimalarials are given to all index household members and 100 neighbours) and increases in the rate at which people seek treatment (treatment seeking is abbreviated as 'TS').

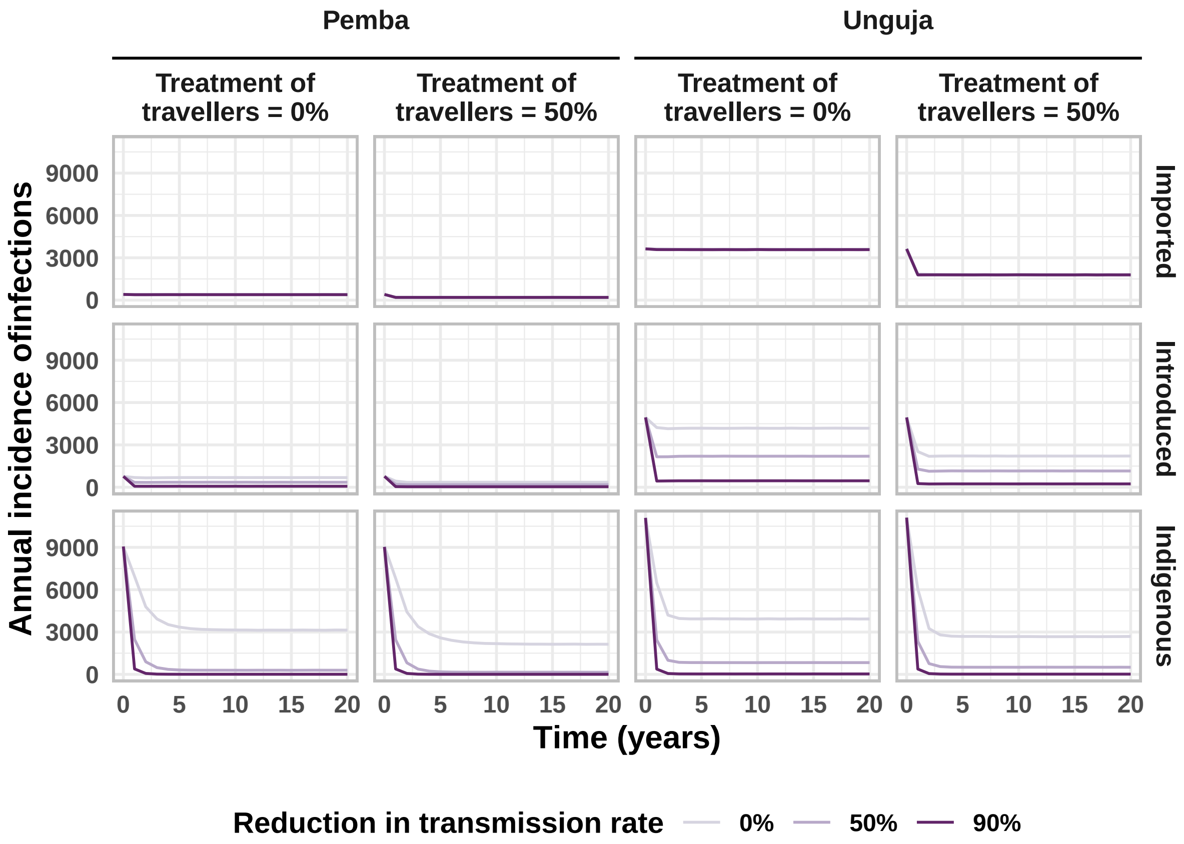


**Figure S12:** Median incidence of infections from 500 stochastic simulations comparing the impact of treating imported cases and reducing the malaria transmission rate on Zanzibar (both Pemba and Unguja). Here, we assume all RCD-related interventions (100% follow up of all cases, treatment of the index household and 100 neighbours in RDA, treatment seeking rate increased to 3 times the baseline rate) are also in effect.

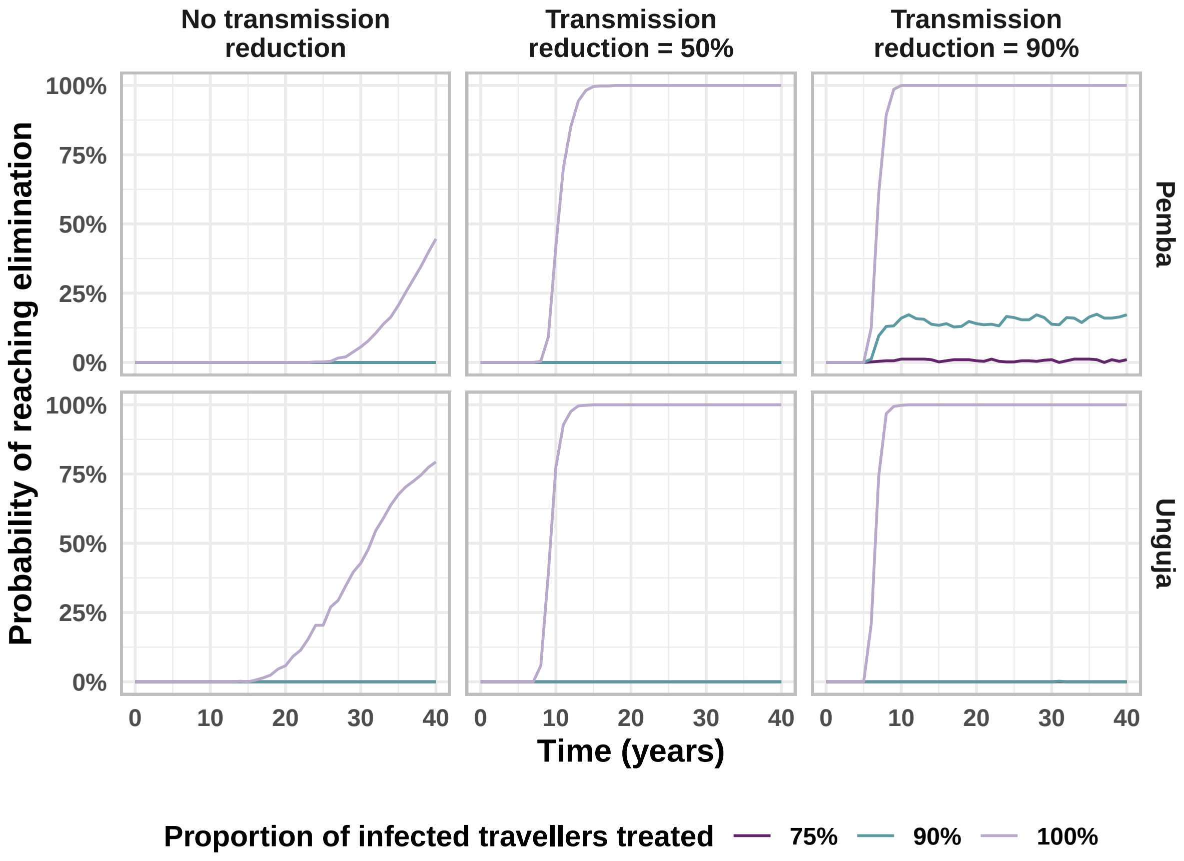


**Figure S13:** Proportion of stochastic simulations reaching elimination (three years with zero indigenous cases), starting from the introduction of interventions. We assume that the maximum values of all RCD-related interventions (100% follow up of all cases, treatment of the index household and 100 neighbours in RDA, treatment seeking rate increased to 3 times the baseline rate) are present and then simulate reducing the malaria transmission rate and treating a proportion of cases imported from mainland Tanzania.

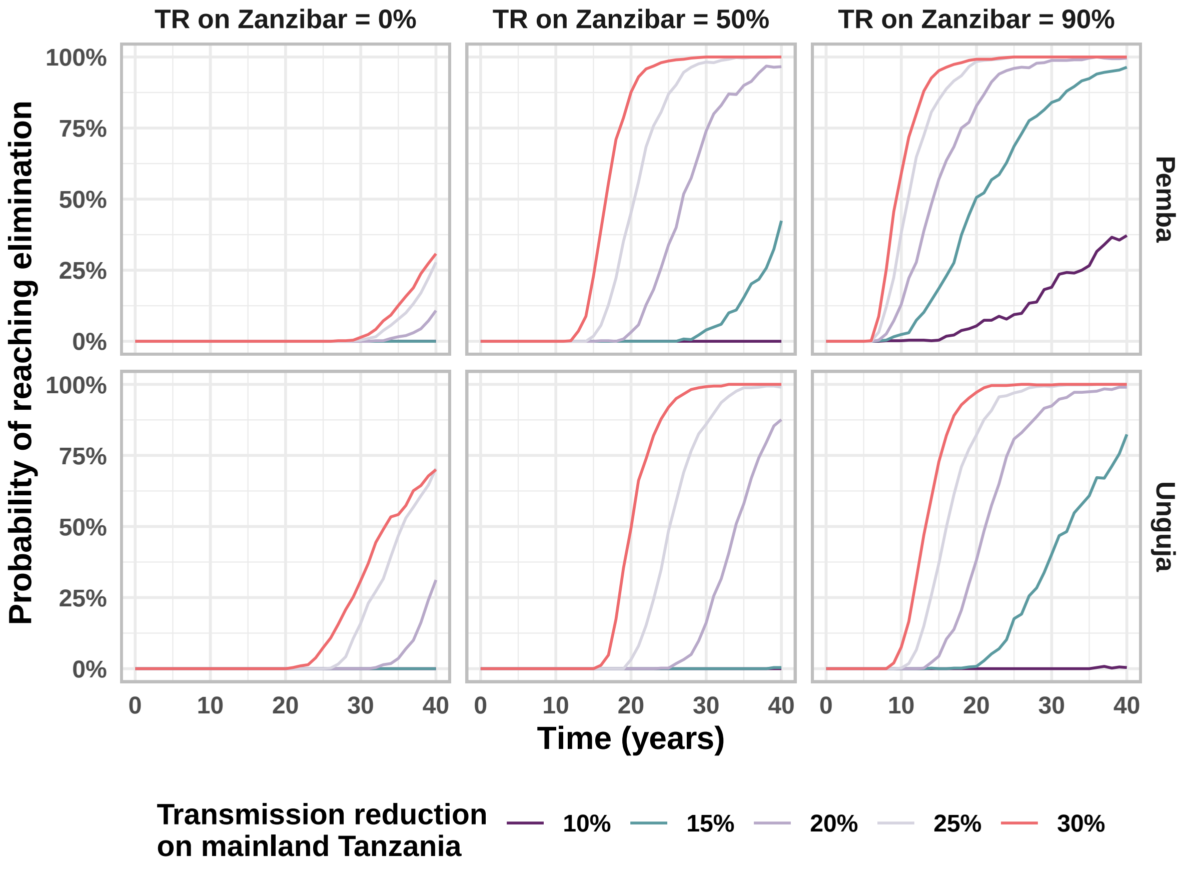


**Figure S14:** Proportion of stochastic simulations reaching elimination (three years with zero indigenous cases), starting from the introduction of interventions. We assume that the maximum values of all RCD-related interventions (100% follow up of all cases, treatment of the index household and 100 neighbours in RDA, treatment seeking rate increased to 3 times the baseline rate) are present and then simulate reducing the malaria transmission rate on both Zanzibar and mainland Tanzania.

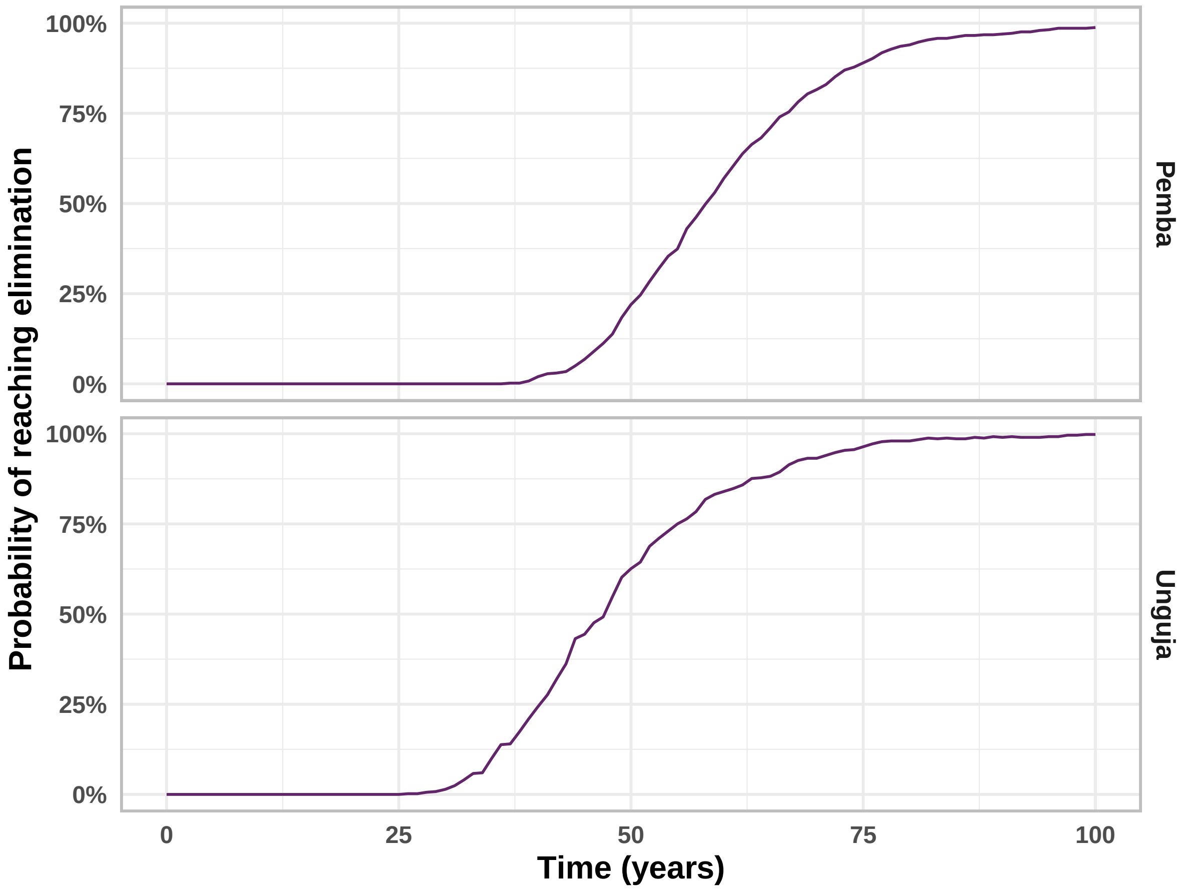


**Figure S15:** Proportion of stochastic simulations reaching elimination (three years with zero indigenous cases) when 100% of infected travellers from mainland Tanzania are treated, starting from year 0. We assume that all other interventions are at baseline values (RCD for 35% of cases arriving at a health facility at the index household level only).
